# Decolonizing WASH Research: Results of a qualitative study and consensus-building process to develop principles for increasing equity in WASH Research

**DOI:** 10.1101/2022.10.16.22281110

**Authors:** J’Anna-Mare Lue, Salamata Bah, Kaelah Grant, Justine Lee, Leila Nzekele, James B Tidwell

**Affiliations:** University of California at Berkeley - Civil and Environmental Engineering; Drexel University - Civil, Environmental, and Architectural Engineering; Drexel University - Computer Science; Drexel University - Psychology and Brain Sciences; World Vision Inc; Drexel University - Public Health

## Abstract

There have long been critiques of colonial legacies influencing global health. In recent years with growing public awareness of unjust systems, a new wave of calls for anti-racist initiatives and decolonization of the sector has emerged. This study examined research inequities in the water, sanitation, and hygiene (WASH) sector, centering the perspectives of researchers from low- and middle-income countries (LMICs), to catalyze collective action in a sub-sector of global health.

Nineteen semi-structured interviews were conducted with researchers of different backgrounds regarding nationality, gender, and research experience. Researchers from eight countries were asked about their experiences and direct observations of discrimination across various stages of the research process. Five interviews were conducted with key WASH research funders to assess perceptions of obstacles faced by LMIC researchers, successes achieved, and challenges faced by these organizations when working towards more equitable research processes within the WASH sector.

The results were analyzed using an emergent framework that categorized experiences based on power differentials and abuse of power; structural barriers due to organizational policies; institutional and individual indifference; othering speech, action, and practices; and context-specific discrimination. The Socio-Ecological Model was also combined with this framework to identify the types of actors and level of coordination needed to address these issues. Respondents were often reluctant to describe actions as discriminatory unless there was clear intent. Researchers who worked in both LMICs and HICs at different career stages were particularly aware of discrimination.

Ensuring pro-equity authorship and funding practices were identified as two significant actions to catalyze change within the sector. Sector-wide efforts must center LMIC voices when identifying research questions, conducting research, and dissemination. Individuals, organizations, and the entire WASH sector must examine how they participate in upholding inequitable systems of power to begin to dismantle the system through the intentional yielding of power and resources.

## Introduction

The linkage between global health and colonialism can be traced from its origins to current global health paradigms and continued partnerships between former colonizing countries and colonized countries (Brown & Bell, 2008; Eichbaum, Adams, Evert, Ho, Semali, & Schalkwyk, 2021). The types of inequities perpetuated by the colonial past of global health determine who has power, resources, and control of the episteme (Fourie, 2018). Multilateral organizations, that set the global health agenda and control much of its funding, have been heavily criticized due to the influence of international politics, resulting in vast inequities in funding allocations and the politicization of health information (Ho, Li, & Whitworth, 2021; Weisz & Nannestad, 2021). These organizations are also considered tools for advancing the economic and political power of their key members who are largely former colonizing countries preserving empires’ control of former colonies (Harrison, 2015; Pearson, 2017).

Additionally, private philanthropy, foundations, and non-governmental organizations also have active roles in perpetuating such inequalities, promoting the “hegemony of neoliberal institutions while reinforcing the ideology of the Western ruling class” (Levich, 2015; Roelofs, 2003).

There are various actors addressing components of Sustainable Development Goal (SDG) 6 to “ensure availability and sustainable management of water and sanitation for all.” Water, sanitation, and hygiene (WASH) is a distinct sub-sector within global health including the global monitoring system of the WHO/UNICEF Joint Monitoring Programme for Water Supply, Sanitation, and Hygiene (JMP), academic conferences, and a distinctive sectoral recognition led by UNICEF within the Humanitarian Cluster system alongside clusters like Food Security and Nutrition. Within the context of WASH, there is an increasing focus on inequity, especially on gendered user experiences of WASH services (Caruso et al., 2021) and imbalances in sectoral leadership (Worsham, Sylvester, Hales, McWilliams, & Luseka, 2021). The history of global health undoubtedly influences the power dynamics found in the WASH landscape. There has been increased scholarly interest in anti-racism and decolonization seemingly due to increased awareness of racialized police violence, especially in the United States, along with the economic and health inequities globally highlighted by the COVID-19 pandemic (Büyüm, Kenney, Koris, Mkumba, & Raveendran, 2020). As a result, longstanding injustices in systems within universities and the scientific literature have been made apparent for all to see (Bhaumik & Jagnoor, 2019; Khan et al., 2019).

Global health research funding is often awarded to or routed through high-income country (HIC) institutions even when research is being conducted within low and middle-income countries (LMICs) (Shumba & Lusambili, 2021). HIC researchers frequently enter LMICs and establish HIC-led and staffed facilities to extract research, which often results in a limited impact on the LMIC’s health systems or research capacity (Eichbaum et al., 2021). Low rates of LMIC authorship of academic publications exemplify the nominal extent to which global health institutions have contributed to capacity-building initiatives in the Global South (Schneider & Maleka, 2018). Given this, it is not surprising that little scholarly research has been conducted to understand barriers and inequalities faced by researchers who are based in or come from LMICs from their perspectives and the resulting impact on systems of knowledge (Luseka, 2020). While there has been well-established criticism of global health work, decolonization is a complex process. This project aimed to investigate inequalities in the WASH sector by centering the experiences of LMIC researchers, examining the root causes of inequity, and exploring feasible strategies for moving toward a more equitable future. The primary objective of this work was to build an anonymized, synthesized base of evidence from which future research, guidance, and initiatives that support LMIC research can be built, ensuring that their contributions are not marginalized, but centered.

## Methods

### Participants

The study participants were LMIC researchers selected with purposive sampling to include males and females and junior and senior researchers in equal proportions. Junior researchers were defined as those who, if they had received a Ph.D., had done so in the past five (5) years. Senior researchers were defined as researchers who had received a Ph.D. at least ten (10) years ago, had obtained funding as lead investigators for at least three (3) projects, and had at least one full-time staff member or student. These definitions were established not to comprehensively cover the types of participants in the research ecosystem, but to intentionally involve those seeking their own funding at an early career stage versus those who were focused on developing and retaining staff and growing a team or organization. The research team reached out to researchers directly and posted on social media (specifically to identify female senior researchers) and requested referrals from participants to establish the final desired sample of 25 interviewees. The sample was not considered to be statistically representative as the project was exploratory—rather, we aimed to gather a breadth of experiences balanced with the desire to take a tractable first step to advance this initiative. We further reached out to representatives of significant funders in the WASH research space to understand their perceptions of such experiences and past as well as ongoing equity initiatives already underway within their organizations.

Participants were informed that the purpose of the study was to gain an understanding of their perspectives and then synthesize and anonymize experiences to then communicate the reality of power imbalances in the sector while minimizing personal risk. Participants were also told that sharing personal or organizational names was not necessary (unless desired, especially to acknowledge positive examples) and that they would not be identified in any way except by gender or career status. Data was stored on a password-protected server, and any identifying details were removed from the transcriptions. The study was approved by the WCG Institutional Review Board (ref: #1-1412585-1; March 17, 2021).

### Reflexivity

The research team represents HIC research institutions and non-governmental organizations which results in an imbalanced power dynamic in the context of this project. However, the majority of the team was comprised of early-career women of color with ties to LMICs in the Caribbean, Southeast Asia, and Africa with education attainment ranging from undergraduate students to master’s degree holders. The first author is a Jamaican woman and a first-year doctoral student in Environmental Engineering. The senior author is an American man with a Ph.D. in Behavioral Science and is an experienced WASH researcher. In consideration of the positionality of the researchers informing the research outcomes, an interview guide was collaboratively formulated by the project team with neutral clarifying and prompting questions to minimize the bias of the research team influencing the study’s findings. When possible two members of the project team conducted interviews, and multiple members of the team reviewed transcripts and participated in coding; the collaborative processes utilized in the research process allowed for dialogue and comparing notes to ensure that research findings reflected the data collected with limited bias from individual researchers.

### Materials

While decolonization and anti-racism may be understudied in a WASH context, the work of critical theorists in the past several decades has created sizable bodies of literature, including well-defined concepts of decolonization, intersectionality, and epistemic violence, which were used as the framing for this study. Kessi’s definition of decolonization as “a political and normative ethic and practice of resistance and intentional undoing – unlearning and dismantling unjust practices, assumptions, and institutions – as well as persistent positive action to create and build alternative spaces and ways of knowing” was followed. (Kessi, Marks, & Ramugondo, 2020). Intersectionality highlights that experiences of discrimination are often not due to a single facet of an individual’s personality and is thus “a lens through which you can see where power comes and collides, where it interlocks and intersects” (Crenshaw, 1989). The concept of epistemic violence emerged from post-colonial feminist critical scholarship (Spivak, 1988); the following definition was utilized “violence against one’s status as a knower; one’s role as a creator and communicator of knowledge… the dismissal of people as credible sources of information, because of our presumptions about them” (Ymous, Spiel, Keyes, Williams, Good, Hornecker, & Bennett, 2020).

The semi-structured interview guide asked about discrimination faced by the LMIC researchers. Given that there were different perceptions of discrimination on the part of participants, we defined discrimination to mean “the unjust making of a distinction on the basis of some attribute about that person by a person or policy that reinforces inequalities.” In facilitating nuanced discussions of the various challenges faced, questions were grouped based on aspects of the research cycle, which included funding acquisition, project execution, and research dissemination, along with more general career advancement.

Finally, experiences with discrimination and mitigating strategies to combat discrimination were elicited. Participants were prompted to voice their own experiences or those they directly observed. Directly observed incidents were included partially to provide anonymity related to describing personal experiences that could be traumatic or potentially harm their careers. Additionally, observations also broadened the potential information gleaned from the sample without compromising the data by including secondhand information. Responses were limited to these situations to avoid including unverified reports in the study while also providing participants with the option to respond that they had no relevant examples to share for any question(s).

### Procedures

Semi-structured interviews were conducted remotely and recorded via Microsoft Teams between March and September 2021. The recordings were then uploaded to an automated transcription website and manually cleaned by a member of the study team. Transcripts were then independently coded by at least two members of the research team using the Dedoose software package (Dedoose, 2018). Codes were derived using an inductive, grounded method, starting by grouping similar challenges expressed by researchers, then arranging these groups into a hierarchy of codes and sub-codes based on relationships observed. The data-derived codes were as follows:

1. power differentials & abuse of power
2. structural barriers due to organizational policies
3. institutional and individual indifference
4. othering speech, action, and practices; and
5. context-specific discrimination.

In addition, a separate set of codes derived from the Social-Ecological Model (Dahlberg & Krug, 2006) were used to assess levels of influence and responsibility for these experiences, both to understand them more fully and to build toward recommendations at each level of the model.

## Results

The sample included a total of 25 participants. There were eleven (44%) early career researchers, eight (32%) senior researchers, and six (24%) funder representatives from five funders, which represented a substantial proportion of the research funding in the sector.

Researchers were from eight different countries; among the 25 participants eleven (44%) identified as male and fourteen as female (56%). Based on iterative analysis of the interviews, the coding structure below was developed (Table 2). With deeply complex and nuanced issues such as discrimination, oppression, and inequity, the recounted experiences described had points of convergence and interrelation as did the codes generated from them. Therefore, many cases were categorized by multiple codes. Due to the exploratory nature of the study and the desire to center participant experiences, substantial detail from these interviews is retained in the analysis below.

**Table 1:**
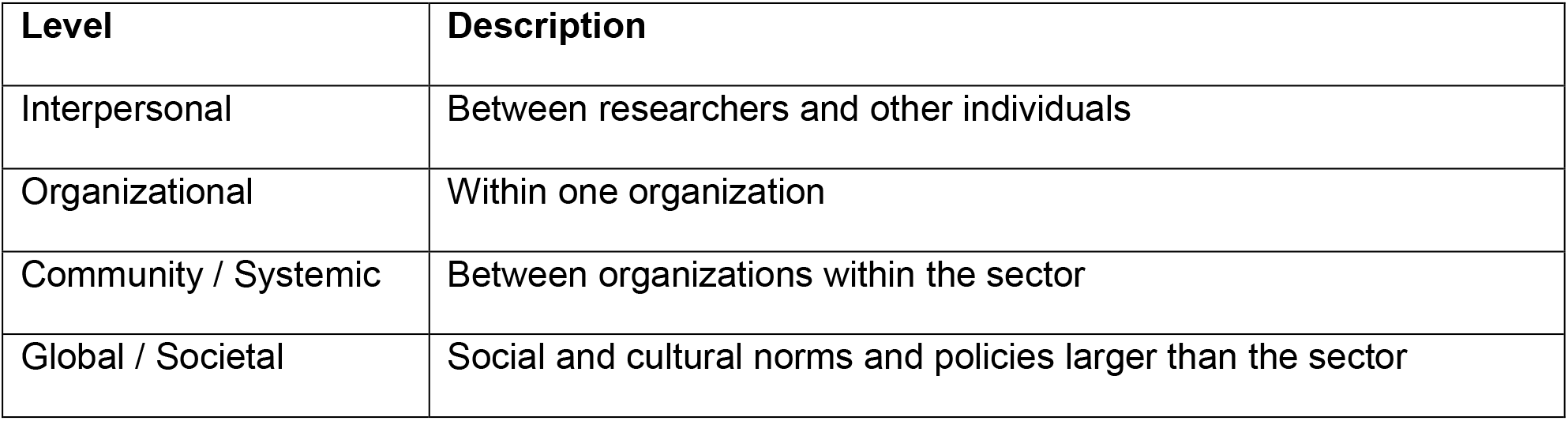
Adapted version of the Social-Ecological Model

**Table 2:** Emergent framework for understanding discrimination and barriers to equity

|  |  |
| --- | --- |
| Power Differentials and Abuse of Power | Disregard for local researcher and community member input |
|  | Lack of LMIC representation on review committees |
|  | Inequitable balance of responsibilities |
|  | Inequitable distribution of authorship and acknowledgement |
|  | Internalized pressure to perform on part of marginalized researchers |
| Structural Barriers due to Organizational Policies | Incompatibilities between funder and recipient systems |
|  | Incompatibilities between research community priorities and local university roles |
|  | Lack of informal networks and insider knowledge by LMIC researchers |
|  | Discriminatory policies related to staff costs, indirects, or other costs |
| Institutional and Individual Indifference | Impacts of English as the predominant language for everyday interactions and knowledge dissemination |
|  | Use of western metrics when evaluating competency |
| Othering Speech, Action, and Practices | From funders or funder representatives |
|  | From journals, publishers, and peer reviewers |
|  | From communities in which research is conducted |
| Context-specific discrimination | Caste-based discrimination |
|  | Tribal discrimination |

### Power Differentials and Abuse of Power

Participants reported both cases of overt abuse of power as well as power differentials that resulted in inequitable and unchallenged assumptions or defaults within the research process. Researchers noted experiences of inequitable distribution of authorship and acknowledgment based on power and privilege. HIC researchers were either awarded prime authorship and desirable leadership roles by default or would demand these positions. In some cases, this was because funding was directly awarded to the HIC institution (sometimes due to funder policies); consequently, the principal investigator, often a senior researcher from a HIC organization, would prioritize their graduate students for prime authorship roles.

In many other cases, LMIC researchers were disproportionately assigned fieldwork and non-technical tasks such as day-to-day data collection, while HIC researchers were able to focus on analysis, interpretation, and writing. This division of labor translated to the HIC researcher maintaining primary ownership over the publication and future presentation opportunities.

This workload imbalance stalled career advancement for LMIC researchers and even affected immigration or visa opportunities. Some LMIC researchers felt that they were meeting the metrics to earn career progression opportunities but were still not considered for advanced roles. One junior female scholar expressed:

> *“I always felt that there was some reason or another that was made up as an excuse for me not to be leading research projects as the [principal investigator] …it’s not always about seniority because some people ended up being promoted and fast-tracked so that they could lead projects on their own. I did receive funding, so I couldn’t believe that that was often cited as the excuse for me not to lead on grants, even when the ideas were mine*.*”*

Funders appeared to be aware of the power imbalances within the WASH field and the resulting discrimination faced by LMIC researchers. One funder representative stated:

> *“[LMIC researchers] do a lot of the work or actually support a lot of the work that’s done in-country level, but [are] not necessarily being included as authors on research manuscripts…that’s a really visible issue: doing the work, but not getting the credit, and not getting the credit impacts your career [and] advancement to publication, right. Publish or perish*.*”*

One senior male researcher also reflected on the impact of restrictive funding calls with set research agendas influencing the type of knowledge production occurring in LMICs and prodding LMIC researchers to conduct research outside of their primary research interests:

> *“[Funding that] is targeted at low-income country settings [is] typically upstream research, fieldwork, data collection, et cetera. I think that is another huge frustration because you as a low-income country researcher [may] want to begin to indulge in more fundamental science or in more discovery, but often funding doesn’t allow [LMIC researchers] to get into those spaces; it very much wants you to go in the field and collect samples*.*”*

There was often a disregard for community members’ and LMIC researchers’ input in research priorities that perpetuated the kinds of research gaps identified and thus the cycle of HIC-centered research priority setting continued. A lack of LMIC reviewers on both funding and publishing committees also amplified the problem.

Seniority was also a factor of power imbalances within the field as well as within organizations. Researchers with less work experience stated that they encountered additional barriers in the research process. One early-career female researcher shared an example of age discrimination when she was selected for a research opportunity and received pushback from older colleagues despite her expertise on the topic. She reflected:

> *“I was wondering why they would be interested in a [WASH] opportunity, if their research area was not WASH? They said, ‘oh this small girl, why are you giving her this opportunity?*’”

Gender inequality was also considered a point of inequitable power distribution. One senior female researcher reflected:

> *“In the sector, I have found myself often as sometimes the only woman in a room of so many men… In some of the research projects, you have like 20 people in a room, and you will just be the only woman there, or you’d be the youngest… There are times when I felt discriminated against, because [as a woman] you have to be not too loud, you have to be quiet*… *Let me also say that I’m a bit passionate in my engagements, but often you get a lot of people feeling that you need to, you know, be a man*.*”*

The disparity of power and resources within the sector leads to an extractive and uni-directional relationship between HIC institutions and communities where research studies are situated. This power dynamic manifests into an internalized pressure to perform on the part of LMIC researchers due to assumptions of incompetency. The internalized pressure aspect was more commonly expressed by women researchers than men. For example, one early-career female researcher stated about the communities in which she was conducting fieldwork:

> *“You really need to prove who you are, really need to work extra hard to show that you can [since] that kind of stereotyping because of your background or where you’re coming from was already there. So, you really need to break these barriers for you to get on*.*”*

Funders reported awareness of these issues and had taken some steps to address them, including mandates and quotas around authorship. However, one funder representative noted internal tensions their own organization faced:

> *“Generally the quality of proposals from Southern institutions were not as strong as those led by Northern institutions—we wanted to favor Southern institutions where we could, but there are obviously different incentives at play and…basic trade-offs around research quality in the short term or building capacity and promoting equity in the longer term*.*”*

Funders also noted that their own organizational policies affected equity in procurement, as described in the following section, but generally reported that the cost of monitoring the internal workings of grantees was too high and that metrics that could be tracked would be very helpful for addressing many aspects of this issue. While some funders did not seem to be able to address the issues of community input, especially where the geographic reach of their programs was extensive, others saw success by focusing in a small number of countries, deeply involving local stakeholders in developing research agendas, and thus shifting power from HIC-based grantees to local stakeholders early in the research process.

### Structural Barriers due to Organizational Policies

Researchers in LMICs face many structural barriers that can be attributed to either formal organizational policies or informal relationships and flows of knowledge. At the most fundamental level, funder and local institutional systems were often fundamentally incompatible. Reporting requirements for funders could often not be met by LMIC-based university accounting systems operations, and LMIC institutions struggled to comply with the tax codes of foreign countries.

Funders were also often restricted either by having to contract directly with institutions in their own country or by legal restrictions on contracting processes between funders and recipients in certain countries. One female early-career researcher decried that:

> *“The funding that I had to apply for had to be [routed] through a [HIC] university. For example, 30% the [HIC] Institute would keep for itself and travel for the project staff from [HIC] to [LMIC]… so, I found it difficult to convince my people in my network to bid for such project. The necessity of having [HIC]-based partners was quite a huge deterrent*.*”*

Participants also reported the sense that even when there weren’t legal restrictions, partnerships with HIC institutions seemed to be expected for grant applications to be successful, with HIC institutions often being the lead on proposals. One senior female researcher stated:

> *“There are two projects I remember*… *we had to use a partner [who is] a research institution based in the donor country as our lead. You can’t just apply if you think your university or institution is qualified, but you have to look for a partner from outside that is based in the donor country in order to be eligible*… *[the funding] is not open [to everyone]*.*”*

Relatedly, the overall organization of local universities was often viewed as incompatible with the research needs of the sector. Often, in this context, local universities placed more emphasis on education than on research and experiential learning even at the most prestigious universities based in LMICs. Furthermore, LMIC-based universities often lacked significant grant-making resources and pipelines of funded Ph.D. and post-doctoral students, which ensure sufficient funding and support research production in HIC universities. As a result, early career researchers were encouraged to go to HIC-based universities, if at all attainable, by all kinds of actors in the research ecosystem. One senior male researcher lamented:

> *“I’ve been a bit torn with the idea of whether I should be promoting students to leave, to go to a Global North university, because they, you know, maybe we’ll get more exposure and more opportunities as a result*.*”*

Beyond official policies and systems, barriers existed due to a lack of “insider knowledge” on the part of LMIC-based researchers. LMIC-based researchers often lacked in-person exposure to funders at international conferences or meetings in funders’ headquarters located in HICs. Differences in exposure were intensified by other forms of discrimination, such as barriers to travel and delays in obtaining visas, such as when acceptance decisions coming from conferences were not given sufficiently far in advance to permit the necessary travel arrangements. One senior female researcher shared:

> *“[The limitation is] our ability to travel and the trust that they do have for people from here, traveling out to certain places. Even if you have evidence in your passport that you’ve traveled to so many places, there’s still so many rules and restrictions*.
>
> *Sometimes the decisions to even attend a particular conference [does] not come five months in advance. Sometimes you are not sure of whether you have a budget for it and then maybe a month to the time they say, we can make a little budget available for you to attend this conference. So quickly, you have to mobilize and get your documents ready and that’s because the sponsorship comes a bit late [and] most of us often rely on sponsorship to travel*.*”*

Several LMIC-origin researchers based at HIC institutions also noted that the experience of HIC institutions internally sharing successful proposals ultimately lead to considerable advantages in these institutions receiving funding.

Finally, inequities related to fees and indirect costs tangibly demonstrated to researchers the disparity in how HIC-based and LMIC-based researchers are valued. LMIC-based researchers were often subject to locally based pay scales determined by their university, whereas HIC-based researchers would often charge higher standard rates or engage through consultancy agreements. On the topic of wages, one senior female researcher stated:

> *“Our salaries in this part of the world are relatively low…So for the time that you’re going to put into that project, ultimately you realize that you are underpaid and yet the bulk of the work is actually going to take place in this part of the world. And I’ve kind of found that always unfair*.*”*

There were also large discrepancies between indirect costs rates. One participant noted that their LMIC institution was only able to allocate 8% of the grant amount for overheads, while HICs were able to charge 40%.

Funder representatives were well aware of the challenges of formal contracting, especially those who were intermediaries funded by a country’s own broader aid budget, and several interviewees were actively working to address those barriers. Success in making such changes slowly seemed to be coming from non-governmental funding agencies, while funders representing government agencies were less optimistic about seeing changes in a reasonable timeline. Few noted approaches underway to share more “insider” knowledge gained by successful applicants over time or had solutions to address costing inequities between institutions. Some expressed awareness of individual organizations active not in WASH, but in adjacent research spaces, especially around developing informal networks to foster collaborative bids. Others mentioned cross-cutting capacity building efforts meant to offset differences in indirect cost rates, but such efforts were still in early stages and there had been few efforts to coordinate or establish norms or guiding principles for these efforts.

### Institutional and Individual Indifference

Institutional and individual indifference to inequity were also manifest in several ways. Passivity towards the challenges experienced by people of differing backgrounds was identified as a recurring theme throughout the interviews. The most significant such obstacle researchers faced was the use of English as the primary language of dissemination.

Researchers whose first language was not English expressed that the language barrier disadvantages them from competing with researchers whose first language is English, regardless of their academic competencies, whether in grant applications, selection to present at conferences, presence of translation services at conferences, or in publication decisions. This indifference also occurred at field level, where meetings with HIC researchers would by default be conducted largely in English with some translation for “field staff,” rather than the other way around.

Additionally, it was observed that researchers from LMICs were being evaluated by metrics originating largely in HICs including formal credentialing and citation of works by others largely outside the context of the research, rather than on the benefit of the research on either its direct subjects or those in similar situations. One early-career female researcher spoke to whose input was considered valuable in research settings, noting that certain projects were not inclusive of younger researchers and community members:

> *“[In] water and sanitation work it’s not only researchers or people with PhDs [who] are involved; it’s different people from different backgrounds, you know, different fields, research assistants and community and different people. And I think sometimes you know, in certain projects, if you’re not highly educated or if you don’t have a certain title, you’re not necessarily included that much*.*”*

This lack of relevant evaluation was seen to invalidate the experiences of the LMIC researchers. The use of such metrics often leads to stagnating or negative career trajectories and personal outcomes regarding position, status, or leadership. One early-career female researcher reflects on stagnancy in career progression experienced as an LMIC researcher:

> *“One of the challenges we are facing as researchers from low- and middle-income countries, the issue of, career progression…The transition from PhD to post-doc then to sort of getting a fixed contract is, is a big challenge because you’ll feel like you are not being given the opportunity*.*”*

Funders sometimes acknowledged that language issues were a challenge, but often limited their focus to trying not to review proposals with consideration for use of “proper English.” There were few noted efforts that would affect their creation or translation before being seen by reviewers or related to activities after grants were awarded. Similarly, alternative impact metrics were sometimes noted as desirable to funders, but there was a gap in identifying appropriate and accessible metrics to use.

### Othering Speech, Action, And Practices

Othering speech, action, and practices refer to the intentional and unintentional discrimination of an individual based on one or more social categories or identities that an individual may hold. Researchers often recounted experiences of feeling othered by funders, their own or other research institutions, organizations involved in publishing, and even the communities in which research takes place.

The power dynamics between funders and researchers often resulted in othering practices that impeded LMIC institutions and researchers’ ability to have a sense of agency. LMIC researchers noted experiences where they were micromanaged by funders and lacked autonomy throughout the research process. One senior male researcher commented:

> *“We have some research funders here right now in my institution who try to interfere into every aspect of what we do. And to an extent, I think that - what if you are going to do this yourself, why don’t you just call yourself a research institution and do it yourself? Sometimes funders overstep their mandate and then really become very insensitive that they’re dealing with a completely separate entity as an institution*.*”*

Throughout the publication process, researchers faced many discriminatory practices. A concern frequently raised was the biased review process for journal publications, where researchers perceived that LMIC institutions without strong partnerships with HIC institutions were not given equal access to publishing opportunities and insider information. One senior male researcher described the practice by saying:

> *“Most research institutes that do international work in Africa have a big brother. They have a big American or European institution that literally runs them. And so, when they write their papers, there’s always a senior author with a particular name that send them. The majority of the work that I do, I do with my teams locally. We’ve, several times, put out what we think are great papers. And we felt that the review that we received was biased. And to be honest we thought that it’s because they don’t expect this level of science coming out of our team, whose names sound African*.*”*

One early-career male researcher reflected on an experience that demonstrated publications preferred HIC credentials and scholars to their LMIC counterparts. The researcher noted that he worked at a HIC institution but serves as a guest lecturer at an LMIC institution when visiting home. He had worked with a student with interesting research questions, and they submitted a paper to a journal with their affiliation being the LMIC institution and it was rejected. The researcher lamented:

> *“We wrote it back with my affiliations as [HIC University] and putting me as the first author in the same journal, it was accepted*.*”*

Beyond othering practices by those within the sector, communities representing society as a whole also speak and act in ways that are discriminatory to researchers. Reported discrimination in the community was largely gender-based, while some reported instances of race and color bias. One senior male researcher noted that his team’s work takes place in a “fiercely patriarchal society” where women researchers do not have the same access to information and resources as their male peers:

> *“Male researchers are able to gain more access to government functionaries than women researchers, particularly if they are traveling alone. And I think it’s important to recognize that particularly women [who] engage in research face unique forms of discrimination that men have no conceivable idea about*.*”*

An early-career female researcher further explained her experience as an engineer, a typically male-dominated career, in the WASH field:

> *“People also think that if you are a woman engineer, you’re not competent enough…I remember when I went somewhere in the rural areas that I’d applied for the job for engineering, I remember I was being interviewed with this old man in the looked me up and down and say, are you sure you’re an engineer?”*

Another early-career researcher recounted discrimination based on skin color when their research team entered communities. He stated the way the team was able to collect the data eventually was by “deploy[ing] other people [from the area] with the electronic survey tools, so they can just be the ones that [research participants] are looking at.”

Funders infrequently mentioned this kind of interpersonal or interorganizational discrimination on their part as a topic of which they were aware or acting to address, though some initiatives underway were noted at specific journals. More general societal discrimination was also infrequently mentioned, and though funders understandably viewed their ability to affect larger cultural values as limited, few micro-level solutions were brought up.

### Context-specific Discrimination

Context-specific discrimination refers to the issues of tribal discrimination, caste-based discrimination, and other forms of discrimination inherent in communities where research is taking place. There is significant intersectionality between these and other types of discrimination previously noted. One senior researcher reflected on the intersectionality of marginalization that women in particular face in LMICs:

> *“When we look at the marginalized, we can go by traditional criteria, which in [an LMIC] context is families will consider either caste, or scheduled tribe, or those considered below the poverty line. But the reality is that discrimination and vulnerability in terms of poverty can transcend castes and economic statuses - a good example of it is women-headed households. Women in our zones will not necessarily fall into the scheduled caste or tribe. And yet can be extremely marginalized. Therefore, as researchers or development practitioners, you need to be open to the fact that discrimination takes multiple forms or marginalization can take multiple forms*.*”*

One early-career female researcher further expounded on gender-based discrimination and tribalism in their country’s context:

> *“I’ve experienced opportunities that [I] have been side-lined on, based on gender and in the part of the country where I’m living. It probably happens in other regions of the country, but we also have a problem of people being discriminated against based on which tribe they come from, which would be more tribalism. In some instances, it is often hard to rise to certain positions if you are not from a particular region, like my current case where I’m working in a region where I’m not from*.*”*

Funders rarely addressed discrimination at such context-specific levels, though some working in more limited geographies did bring up the issue, especially noting the non-homogeneity of LMICs and the idea that “local institutions” may be viewed as outsiders in some locations within the country.

## Discussion

This study sought to highlight the experiences of LMIC researchers as a basis for establishing more equitable approaches to research and the generation of knowledge around WASH as a microcosm of global health. The nature of the organization and functioning of the WASH sector, the establishment of global targets primarily focused on minimum standards, and international collaborations between HIC and LMIC institutions and researchers, lends itself to power disparities at the personal and inter-organizational level.

Power differentials encourage inequitable research partnerships, conflicting research priorities, and disparities in funding, authorship, and recognition. (Shumba & Lusambili, 2021; Faure, Munung, Ntusi, Pratt, & de Vries, 2021) The absence of LMIC representation within funding organizations and review committees perpetuates inequality within the sector. Furthermore, incompatibilities across different levels and systems within the research ecosystem result in substantial discrimination, but a lack of understanding or prioritization by powerful actors means such incompatibilities persist. Internalized pressures encountered by LMIC researchers, lack of informal networks, and discriminatory metrics hinder progression towards more just frameworks and practices in WASH research.

### Limitations

We note at the outset of the discussion several key limitations in our findings. First, although participants were diverse in terms of country of origin, there is no claim of representativeness either within countries with a wide range of contexts and experiences or across the large number of settings where WASH research is conducted. This limitation is attributed to both the size of the sample and the method of recruitment via existing relationships and referrals. In particular, context-specific sources of discrimination, such as the caste system in India, are certainly underexplored. Second, and a more fundamental challenge in studying this topic, is that the selection of researchers active in the field does not capture the experiences of people who were not able to enter or continue in the field, and substantially different kinds of discrimination may have led to some of the challenges those people faced. Third, many LMIC researchers interviewed noted that despite feeling frustrated by particular experiences, they did not fully become aware of the discriminatory nature of the incidents until further reflection afterward, often when moving to another context for instance working in a HIC. There was also hesitancy in labeling a policy or action discriminatory. The importance of understanding experiences that are inevitably colored by the individual’s perspective makes it challenging to objectively identify the relative and absolute scale of different forms of discrimination. Finally, although representation based on national origin, gender, and career stage was ensured, other significant categories of marginalization such as sexual orientation and disability were challenging to observe without participants potentially risking job security, criminalization, and further discrimination.

### From The Margins to Center – Imagining Equitable WASH Research

The WASH sector primarily provides services to vulnerable communities predominantly in the Global South. Research equity begins with the acknowledgment of marginalization and systems of inequities by all actors, including LMIC and HIC research institutions, funders, governments, multilateral organizations, and scientific journals. People in the WASH sector speak extensively of the inequities in access to WASH infrastructure and knowledge, and the human right of access to clean drinking water and adequate sanitation. Several individuals have also penned articles both explaining the state of the WASH sector and calling for action to begin a decolonization process. However, peer-reviewed literature on decolonizing WASH research and larger knowledge generation practices is sparse. We, therefore, draw upon generations of scholars who have produced scholarship within the areas of post-colonial and different ethnic studies, critical theory, Black and global feminisms, and queer studies of which strategies and frameworks of justice and decolonization can be utilized as a basis for improving equity in the context of WASH. We organized recommendations generated by this process at four levels based on the Social-ecological Model of Behavior (Dahlberg & Krug, 2006), including interpersonal, organizational, community/systemic, and global/societal issues. We note that such a list of interpersonal or organizational steps could be resolved by unilateral action, but that many broader changes require collective action and enforcement of norms that disincentivize a “race to the bottom” when it comes to policies related to LMIC researchers. We, therefore, discuss a few such key results where collective action may catalyze change.

First, LMIC authorship has been posited as a critical part of a more equitable research process. Authorship and other leadership opportunities for LMIC researchers allow access to more options for career advancement and agency in the research process. (Urassa, 2021) Our study also found that epistemic violence is prevalent in the WASH research arena, as there are common assumptions of LMIC researchers’ competence based on nationality, command of the English language, and other western metrics. The WASH sector must ask whether WASH is a space where the LMIC researchers are empowered to share histories, context, and indigenous ways of knowing and to produce academic knowledge. Epistemic violence must be addressed to achieve improved equity in research authorship and leadership. (Fourie, 2018) As those that hold the power, journals should develop and enforce standards for publications based on research set within LMIC contexts. However, given that policies at the level of the publication cannot be overly strict (e.g., absolutely requiring that the first author is from the country where the research took place), funders and research institutions must establish and publicize metrics related to equitable publishing practices.

Second, based on the findings of discriminatory funding and publishing practices that place emphasis on collaboration with HIC institutions and have skewed funding allocations between HIC institutions and LMIC ones, there is a need to further promote the development of research capacity in LMICs. Although funding calls that recommend or require collaboration with a HIC institution may intend to nurture international knowledge exchange, such assumptions may affect more harm than good by inhibiting LMIC research career advancement and research autonomy. Additionally, if not adequately resourced, capacity building cannot be effective. Direct and indirect research funding is vital for increased justice in WASH research.

Global health has been charged with neo-colonialism in thousands of scholarly articles over the years, including by David Beran and co-authors (Beran, Byass, Gbakima, Kahn, Sankoh, Tollman, Witham, & Davies, 2017) who state, “The neo-colonialism of global health has muted the local voice, and a lack of long-term investment in infrastructure has made institutes and researchers in many LMICs ill-equipped to find local solutions to local problems.” While current practices in WASH have advanced the field to where it currently stands, new approaches to conducting and funding research and building research capacity must be implemented to ensure that there is equity within the WASH ecosystem.

Research institutions and funders should establish agreed-upon standards for budgetary allocation levels, both at the institutional and project level, for capacity-building efforts. Taken together, collective action towards equity in funding and authorship will engender the processes needed to drive other collective changes and spur individuals and organizations to make changes that require only their own internal processes or decision-making as well.

bell hooks, prominent Black feminist scholar, stated that “to be in the margin is to be part of the whole but outside the main body.” (hooks, 1984) LMIC communities, researchers, and institutions are in the margins of their own experiences with WASH in research.

Decolonization and improving research equity must center these stakeholders, bringing them to the forefront to tell their own stories, set research agendas, and gain credit when due.

Centering in this context requires the “yielding of power” and resources to stakeholders who are of less privileged identities due to social and geopolitical factors. (Eichbaum et al., 2021) Collective action to support equitable authorship, research funding, and capacity-building programs, can begin to unlock the power and bring justice to those too long marginalized.

## Conclusion

As the question of decolonization and equity in WASH research and practice continues to be raised, all actors involved in the research process must take action toward meaningful solutions. This study was intended to delve into the specific case of WASH research equity as a microcosm of global health and provide a basis for financial and epistemic investment in a more equitable WASH research landscape. LMIC researchers’ and funders’ own experiences indicate that the colonization of WASH knowledge is widespread.

Acknowledging the experiences of LMIC actors is an important first step to achieving equity, but the roles of individuals, and organizations who build and maintain oppressive systems within global health, and by extension WASH, must be examined and then dismantled.

Without an intentional yielding of power, there can be no justice in WASH research and practice.

## Data Availability

All data produced in the present study are available upon reasonable request to the authors.

## Conflicts of Interest

None reported.

## Funding

This project was funded by World Vision.

## Notes

### Competing Interest Statement

The authors have declared no competing interest.

### Funding Statement

This study was funded by World Vision.

### Author Declarations

The study was exempted by the WCG Institutional Review Board (ref: #1-1412585-1; March 17, 2021).

